# Stigma, perception, and lived realities: An ethnographic study of cutaneous leishmaniasis in rural Ethiopia

**DOI:** 10.1101/2025.08.01.25332707

**Authors:** Binega Haileselassie, Zenawi Zerihun, Afeworki Mulugeta

## Abstract

**Background:** Cutaneous Leishmaniasis (CL) is a neglected tropical disease with profound physical, psychological, and social consequences, particularly in low-resource settings. In rural Ethiopia, Tigrai, CL remains poorly understood and heavily stigmatized, shaped by deeply rooted sociocultural perceptions that interpret the disease through supernatural, moral, and spiritual frameworks. These beliefs affect how affected individuals are perceived and treated within their communities, often resulting in social exclusion and delays in care.

**Methodology/principal findings:** This ethnographic study was conducted in rural districts of northern Ethiopia, Tigrai, using ethnographic field observations, in-depth interviews, and focus group discussions with affected individuals, caregivers, Health Extension Workers (HEWs), and Community Advisory Group (CAG) members. Findings reveal that CL is widely perceived as a condition linked to divine punishment, impurity, or ancestral curses. These beliefs contribute to stigma, emotional distress, and restricted social participation, especially among women and children. Structural barriers including economic hardship, limited access to biomedical care, and poor health literacy further exacerbate the disease burden. While HEWs and CAGs attempt to address misconceptions and promote treatment uptake, their efforts are constrained by systemic resource limitations and community resistance.

**Conclusions/significance:** CL in rural Ethiopia, Tigrai, is not only a biomedical issue but also a socially constructed and culturally embedded affliction. Addressing the disease requires a holistic public health response that integrates ethnographic insights, respects local belief systems, and prioritizes stigma reduction and community engagement. These findings contribute to a growing body of literature emphasizing the need for culturally sensitive, equity-driven approaches in global NTD programming.

**Author summary:** CL is a skin disease caused by parasites and transmitted by sandflies. While it may not cause death, it often leads to visible skin scars that have serious emotional, social, and economic impacts especially in rural areas with limited access to healthcare. In this study, we explored how people living in rural Ethiopia, Tigrai region, experience and understand CL. Through interviews, focus group discussions, and field observations, we found that many community members believe CL is caused by supernatural forces, divine punishment, or moral wrongdoing. These beliefs cause people with CL to be stigmatized and socially excluded, particularly women and children. People often turn to traditional remedies instead of modern treatment, which delays recovery. We also learned that community health workers try to educate people and support treatment, but they face many challenges. Our study shows that treating CL should involve more than just medicine it must include efforts to reduce stigma, engage communities, and understand local beliefs. These findings can help improve public health programs for CL and similar neglected diseases.

## Introduction and background

Cutaneious leishmaniasis, classified as a neglected tropical disease (NTD), constitutes a significant public health concern across multiple regions globally, including Ethiopia. Despite its relatively low mortality, CL causes profound morbidity, both physically and socially, with sufferers often bearing long-lasting scars visible markers that carry social weight far beyond the biological infection. The physical disfigurement caused by the disease intersects with deeply held cultural, spiritual, and social beliefs that frame the illness not merely as a medical concern, but as a moral or spiritual blemish. As a result, individuals affected by CL often encounter stigma, social isolation, and discrimination, which, in turn, negatively impact their psychological well-being and health-seeking behavior [1, 2].

Historically, CL has been prevalent in marginalized and underserved communities where access to quality healthcare, accurate information, and social protection mechanisms are limited [3, 2]. In such settings, traditional beliefs frequently dominate the health discourse, often framing CL as a punishment from supernatural forces, a curse, or a result of moral failings or impurity.

These beliefs not only stigmatize the disease but also reinforce community-wide behaviors that delay early detection and treatment. In the Ethiopian context, particularly in rural areas, these challenges are compounded by poverty, limited health infrastructure, gender-based disparities, and poor awareness at both individual and community levels [4].

The neglect of CL in national and global health agendas has also perpetuated the disease’s invisibility and the lived struggles of those who endure it. While biomedical research has addressed the parasitological and epidemiological aspects of CL [5], less attention has been paid to the social suffering, local explanatory models of illness, and coping strategies employed by affected individuals and communities. As [6] asserts, diseases of poverty are not just biological phenomena but are shaped and sustained by political, economic, and social inequalities a perspective echoed by anthropologists and public health scholars who emphasize the importance of qualitative and ethnographic approaches in understanding the full scope of neglected diseases [7, 8].

This paper is situated within this interdisciplinary gap. It aims to provide a nuanced, context-specific understanding of the stigma, perception, and lived realities of people affected by CL in selected rural districts of northern Ethiopia. The ethnographic fieldwork, conducted over several months, incorporates the voices of those often excluded from policy conversations: patients, caregivers, HEWs, and CAG members. Their insights not only reveal the hidden burden of the disease but also highlight culturally rooted barriers to healthcare access and acceptance. This study, therefore, contributes to the body of knowledge advocating for community-centered, culturally informed public health interventions.

CL remains one of the most overlooked tropical diseases, particularly in low-resource settings where it is endemic. While biomedical research has advanced the understanding of the disease’s parasitic causes and therapeutic options, there is a persistent gap in understanding the sociocultural and psychological dimensions of CL, especially in endemic rural communities [3, 2]. In Ethiopia, despite its inclusion in public health initiatives, CL continues to be inadequately addressed in terms of stigma reduction, mental health impacts, and culturally appropriate education and outreach [9, 8].

The prevailing body of literature on CL is dominated by epidemiological and clinical studies, often neglecting the experiential realities of those living with the disease. Few studies have systematically explored how traditional belief systems, spiritual interpretations, or social hierarchies affect how CL is perceived, diagnosed, or treated at the community level. Even fewer have examined the gendered nature of stigma, its impact on educational opportunities, employment, or marriage, and how these social consequences are negotiated by individuals and families [1, 4].

Moreover, the narratives and emotional burdens of caregivers and community-level health actors, such as HEWs and CAG members, are seldom represented in health discourse, policy, or programming. This exclusion not only weakens intervention strategies but also limits the sustainability of health initiatives due to lack of contextual adaptation [7]. As such, there exists a critical need to integrate ethnographic insights into the understanding of CL, especially in regions where the disease is deeply intertwined with socio-spiritual narratives and structural barriers to healthcare.

This study seeks to bridge this research gap by providing an in-depth, grounded account of the lived experiences of individuals affected by CL in northern Ethiopia. Through qualitative ethnographic methods including participant observation, interactive interviews, and focus group discussions it captures the complex interplay between stigma, local beliefs, emotional suffering, and institutional response. This work responds to calls for more inclusive and culturally relevant research approaches in tackling NTDs and seeks to inform both local and global health strategies with contextually rich evidence. Therefore, this essay explores and analyze the sociocultural perceptions, stigma, and lived experiences associated with CL in rural Ethiopian communities, with the aim of informing culturally responsive public health strategies.

## Method

### Study design

This research employed a qualitative ethnographic design aimed at generating deep, contextualized understanding of the lived experiences of individuals affected by CL.

Ethnography, as both a methodological orientation and data collection strategy, was chosen for its strength in uncovering cultural meanings, social practices, and the embodied dimensions of health and illness [10, 11]. The approach allowed for immersive observation and extended interaction with participants in their natural settings, offering nuanced insights that are often missed in quantitative surveys or clinical studies.

### Study setting

The fieldwork was conducted in three rural districts of Northern Ethiopia: Degua Tembien, Sewha Saese, and Emba Alaje. These areas were purposively selected based on their known endemicity of CL, accessibility, and community willingness to participate. The districts are primarily agrarian and characterized by limited infrastructure, high reliance on traditional beliefs, and strong religious and cultural institutions, all of which have implications for health behavior and disease perception.

### Study population and sampling

The study population included individuals diagnosed with CL (either with active lesions or healed scars), caregivers of affected persons, HEWs, CAG members, traditional healers, and community elders. A purposive and snowball sampling strategy was employed to recruit a diverse cross-section of participants. Inclusion criteria required that participants be residents of the area, have either direct or indirect experience with CL, and consent to being involved in the study. A total of 42 in-depth interviews and 6 focus group discussions were conducted with approximately 64 participants. Sampling continued until thematic saturation was reached that is, when new data no longer contributed additional insights.

### Data collection methods

Data were collected over a six-month period using a variety of ethnographic tools:

**Participant observation.** The researcher spent extended periods in the study communities, attending local events, shadowing HEWs, and observing everyday interactions in households, farms, markets, and health posts. Field notes captured verbal and non-verbal expressions of stigma, social distancing behaviors, and coping strategies.

**In-depth interviews.** Semi-structured interviews were conducted with affected individuals, caregivers, health workers, and key informants. Interviews explored personal narratives of illness, beliefs about disease causation and healing, experiences of discrimination, emotional distress, and health-seeking behavior.

**Focus Group Discussions (FGDs).** FGDs were held with community members and health stakeholders to elicit collective views on CL, community norms, and existing intervention challenges.

All interviews and discussions were conducted in Tigrinya, the local language, and later translated into English by the researcher and a bilingual assistant. Informed consent (verbal and written) was obtained from all participants.

### Data analysis

A thematic content analysis approach was employed. Transcribed data were coded both inductively and deductively using NVivo software. The inductive process allowed themes to emerge organically from the narratives, while deductive codes were informed by the literature on stigma, health-seeking behavior, and community engagement. Key themes identified included: social exclusion, explanatory models of CL, gendered stigma, trust in health systems, and traditional healing practices.

Data triangulation across interviews, observations, and FGDs enhanced the validity of findings. Reflexivity was maintained throughout by documenting the researcher’s positionality, potential biases, and interactions with participants.

### Ethical considerations

This research forms part of the ECLIPSE programme, funded by the National Institute for Health and Care Research in the United Kingdom. Ethical approval for the ECLIPSE programme was granted by the Health Research Ethics Review Committee (Ref. ERC/1793/2020) in Ethiopia and the Faculty of Medicine and Health Sciences Research Ethics Committee at Keele University, United Kingdom (Ref. MH-200123). We also obtained a support letter from Tigray Health Bureau and the College of Health Sciences at Mekelle University, Ethiopia. Participants were assured of confidentiality, voluntary participation, and their right to withdraw at any point. Pseudonyms were used to protect participant identities. Sensitive interviews, particularly those dealing with stigmatization and emotional distress, were conducted with empathy and followed by debriefing where needed.

### Limitations

While the ethnographic method offered rich, grounded insights, several limitations were acknowledged. These include potential language translation bias, limited generalizability beyond the study communities, and the possibility of social desirability bias in responses about stigma. Additionally, due to security issues in the region at the time of fieldwork, some locations and groups could not be accessed as originally planned

## Result

The ethnographic fieldwork revealed a complex web of community beliefs, psychosocial experiences, and systemic challenges surrounding CL. Thematic analysis yielded six major themes: (i) Misconceptions and cultural interpretations of CL, (ii) Psychosocial suffering and emotional distress, (iii) Gendered stigma and social exclusion, (iv) Barriers to modern healthcare, (v) Role of HEWs and CAGs in awareness creation, and (vi) Work and school impairments linked to disease burden.

### Misconceptions and cultural interpretations of CL

A majority of community members associated CL with supernatural causes such as divine punishment, ancestral curses, or exposure to spirits. An elderly man in Emba Alaje explained, “We used to think CL came from angering the spirits or being sinful. It is not a normal illness.” This statement underscores the entrenchment of cultural and religious belief systems in explaining illness, which often supersede biomedical understandings. The disease was also commonly seen as a manifestation of impurity, particularly when lesions were visible, reinforcing social exclusion.

> *People say I have this disease because I did something wrong, that I offended the spirits. Some believe it came from a moth or a curse. They don’t touch me, they don’t visit. Even my neighbors stopped greeting me. My family took me to holy water instead of the health center because they thought prayer would help more than medicine (P03, Alaje).*

This narrative encapsulates how CL is culturally constructed as a morally charged condition, deeply entangled with spiritual and supernatural explanations. The respondent’s experience of social avoidance and religious-based care-seeking illustrates how stigma, misinformation, and communal beliefs override biomedical logic. The loss of social ties and preference for holy remedies reflect a health system dislocated from the community’s worldview making healing not just a medical challenge, but a social and symbolic one.

Some believed CL could be transmitted through contact with bats, moths (*black gizwa*), or the urine of infected animals highlighting a critical gap in public biomedical knowledge.

These perceptions have tangible social consequences. As one caregiver stated, “People think my child’s condition is a curse. They avoid us, and it hurts deeply.” This illustrates how stigma stemming from misinformation can alienate families and exacerbate emotional suffering. The prioritization of traditional remedies such as holy water and herbal treatments, often at the expense of modern medical options, further reflects the community’s reliance on culturally sanctioned healing practices.

### Psychosocial suffering and emotional distress

Participants expressed profound emotional distress, including anxiety, self-stigmatization, and diminished self-worth. A young woman with a facial lesion shared, “I always feel like people are staring at me. It makes me anxious. I don’t want to go to church or to the market anymore.” This quote reveals the internalization of stigma, leading to social withdrawal and emotional isolation. The feeling of being scrutinized in public spaces resulted in affected individuals limiting their participation in communal and religious life.

> *When I walk through the village, I feel their eyes on me. It’s like everyone is watching, judging. I stopped going to the market, and I haven’t been to church in months. I just stay at home now it feels safer not to be seen (P41, Degua Tembien).*

Caregivers also bore significant emotional burdens. One mother expressed, “Seeing my child suffer breaks my heart. I worry not just about the disease, but how society will treat him in the future.” Her words highlight how stigma extends beyond the afflicted to encompass the family, creating layers of psychological strain. The uncertainty surrounding future discrimination compounds the immediate stress of caregiving.

> *Every time I look at my child, I feel sorrow not only because of his wound, but because I know how people talk. They say it’s a curse, and I wonder what this will mean for him when he grows up. I’m scared of the future they’ve already written for him. (P36, Subha Saese).*

These accounts reflect the layered emotional toll CL inflicts on both the afflicted and their caregivers. The young woman’s retreat from public spaces reveals how stigma becomes internalized, eroding self-worth and severing social participation. Meanwhile, the other narrative demonstrates how stigma anticipates and shapes imagined futures, projecting social suffering forward in time. Together, they highlight the psychosocial dimensions of CL that go beyond physical symptoms, revealing how the disease not only scars the skin, but silently reshapes identity, kinship, and community belonging.

### Gendered stigma and social exclusion

Stigmatization disproportionately affected women and girls, particularly in relation to societal roles and marital prospects. A focus group participant remarked, “Even if a girl is beautiful, if she has a lesion, people say she is cursed. No man will come for her.” This comment reflects how visible signs of illness are interpreted through a gendered lens, where female value is often tied to physical appearance and social desirability. A young women remarked,

> *They say no one wants a girl with a scar. Even if she is kind or clever, people only see the mark on her face. Parents whisper about her like she’s not worthy of marriage. It’s as if the lesion erased everything else about her (P16, Degua Tembien).*

While boys also experienced stigma, their social mobility remained relatively less constrained. The intersection of gender and health thus further marginalized females, limiting their opportunities and reinforcing structural inequalities. Participants often talk of the gendered perspective of CL. A father stated this:

> *My daughter cried when she heard a neighbor say, ‘She’s ruined now.’ She’s just a child, but already they’ve decided her future. If she were a boy, they wouldn’t speak that way. It’s different for girls everyone watches, everyone judges (P 27, Alaje).*

These testimonies unveil the deeply gendered nature of CL stigma, where visible lesions are read not just as signs of disease, but as blemishes on a woman’s social value and future potential. Female identity becomes tightly policed through aesthetics and marriageability, with public narratives reducing girls to their physical appearance. In contrast, boys enjoy greater leniency, their worth tied less to bodily “purity.” This asymmetry reinforces patriarchal norms, situating disease not only within the body but within structures of inequality that regulate femininity, honor, and visibility.

### Barriers to modern healthcare

Participants identified several barriers to accessing biomedical care, including prolonged wait times, cost, and perceived ineffectiveness. One woman recounted her experience at Ayder Hospital: “They told me to come back after three months. I have children to feed. I couldn’t afford to wait.” This statement illustrates how economic precarity intersects with institutional delays, making modern healthcare inaccessible to many.

> *When I reached the clinic, they gave me an appointment for two months later. I walked three hours to get there, leaving my children behind. I don’t have money to keep coming and going like that, so I just stopped trying (P22, Subha Saese).*

This account illustrates how logistical and economic burdens compound to deter access to biomedical care. The intersection of rural geography, unpaid caregiving responsibilities, and delayed treatment scheduling not only weakens trust in health institutions but effectively excludes vulnerable populations from receiving timely interventions.

> *At the private clinic, they gave me an injection that made the wound worse. I spent all the money I had and felt more pain than before. After that, my mother took me to a holy site instead. At least there, I felt some hope (P11, Alaje).*

Here, the respondent’s disillusionment with private-sector care reveals how high costs and poor outcomes drive individuals toward spiritual healing practices. The preference for sacred sites over clinics reflects both a crisis of medical confidence and a reliance on culturally meaningful sources of healing, highlighting the complex interplay between perceived efficacy, affordability, and faith.

In addition, some reported that treatments from private clinics were both costly and counterproductive. These perceptions, combined with a prevailing belief in spiritual remedies, delayed treatment initiation and reduced adherence. The health system’s limitations contributed to continued reliance on traditional and informal care.

### Role of HEWs and CAGs in awareness creation

Despite the challenges, HEW) and CAGs played pivotal roles in community education. A HEW explained, “We try to educate people, but the myths are deeply rooted. It’s a constant battle to change their minds.” This reflects the entrenched nature of cultural beliefs and the perseverance required by frontline health workers.

> *Some families still believe CL comes from spirits or curses, so when I talk about sandflies or parasites, they look at me like I’m speaking another language. I’ve gone back to the same house three or four times just to be heard. It’s not easy, but I can’t give up on them (HEW 01, Degua Tembien).*

This quote captures the persistence and emotional labor involved in frontline health education. The HEW’s experience underscores the epistemological divide between biomedical science and local belief systems, requiring not only medical knowledge but cultural fluency and trust-building. Her repeated visits signal how public health work in such context’s hinges on long-term engagement, not quick fixes.

CAG members helped bridge the gap between biomedical knowledge and traditional understanding. As one CAG leader emphasized, “Our group works hard to support families and educate the community. It’s slow progress, but we are making a difference.” Their commitment to grassroots advocacy and culturally respectful dialogue was essential in fostering incremental shifts in community attitudes.

> *When people don’t listen to the health workers, we step in. We use stories and examples from the community they trust us more because we are part of them. I’ve seen people slowly change, not because they were forced, but because they felt understood (CAG 03, Subha Saese).*

This account illustrates the strategic role of CAGs as cultural mediators who translate health messages into familiar narratives. Their embeddedness within the community fosters trust and gradual attitudinal change, emphasizing that effective health communication must resonate emotionally and socially, not just scientifically. Their work reflects a model of health promotion rooted in dialogue rather than authority.

### Work and school impairments

The functional impacts of CL were significant. Individuals with lesions described daily limitations, particularly in agricultural labor. A woman remarked, “When I bend down to weed, the lesion throbs. I rest more than I work.” This statement highlights the disabling pain and its effect on productivity, especially among subsistence farmers.

Children with CL faced educational setbacks due to stigma and physical discomfort. A ninth-grade student shared, “When I write or read, the pain gets worse. I had to miss exams and I fell behind.” Such testimonies underscore how disease burden can hinder educational attainment, affecting long-term life opportunities and reinforcing cycles of poverty. A wone farmer remarked:

> *Before I got this sore, I could carry water, farm, and cook for my family. Now, even bending down sends sharp pain through my leg. I stop often, and my husband complains that I am lazy. But I’m not lazy, I’m just in pain (P 24, Alaje).*

This testimony reveals how CL disrupts not only physical labor but also social relationships and gendered expectations of productivity. In agrarian households, where women are central to both food production and caregiving, such impairments carry a dual burden: bodily limitation and moral judgment. The respondent’s experience of being perceived as “lazy” reflects broader cultural narratives that conflate illness with personal failure, especially among women.

This dynamic contributes to internalized stigma and deteriorates intra-household support, compounding the isolation of those affected. In contexts where physical contribution is central to social value, CL marks individuals not just as ill, but as burdensome. People with CL talks frequently about this and one participant highlighted:

> *My hand hurts when I write, and sometimes blood stains my exercise book. When I miss school, my classmates move forward, and I stay behind. I want to be a teacher someday, but I feel like that dream is running away from me (P 34, Subha Saese).*

The participant reflection highlights the subtle yet profound educational consequences of CL, particularly for children who are navigating both academic expectations and the psychosocial weight of visible disfigurement. Pain-related absenteeism and embarrassment undermine not only classroom participation but the child’s long-term sense of academic identity and hope. Dreams of upward mobility such as becoming a teacher become eclipsed by a mounting sense of inevitability and exclusion. His testimony underscores how neglected tropical diseases like CL not only disrupt immediate learning but also erode the foundational belief in a possible future, reinforcing cycles of intergenerational poverty and social stagnation. Another participant also revealed this as follow:

> *They said I shouldn’t touch the tools because of my wound. I begged them, but they said customers might complain. Now I sit outside the market and wait for someone to ask for help, but no one ever does (P 38, Degua Tembien).*

This reflects the economic marginalization of CL sufferers within informal labor markets, where bodily integrity and perceived hygiene are implicitly tied to employability. The visible lesion becomes a form of symbolic contamination, marking the individual as unfit for both physical work and social trust. Beyond job loss, the young man experiences a kind of existential exclusion waiting daily in public spaces not only for work but for recognition. His social invisibility is emblematic of how CL transforms from a biomedical condition into a form of structural abandonment, with implications for livelihood, self-worth, and communal participation. The quote illustrates how stigma becomes institutionalized in labor relations, deepening socioeconomic precarity.

## Discussion

The ethnographic findings presented on Cutaneous Leishmaniasis (CL) provide a rich, multi- dimensional portrayal of the lived experiences and sociocultural interpretations surrounding the disease. In synthesizing these findings within the broader scholarly discourse, both resonance and critical dissonance emerge, offering nuanced insights into how neglected tropical diseases are experienced at the intersection of health, culture, and social structure.

A recurring theme in global health literature is the disjuncture between biomedical and local explanatory models of disease. This study confirms that community members interpret CL through spiritual and moral frameworks viewing it as divine punishment or ancestral displeasure. [12] similarly documented persistent supernatural beliefs among CL patients in Ethiopia, highlighting the challenge of bridging traditional epistemologies with biomedical health literacy. [13] further underscore misconceptions about transmission such as via animals or bats and the reliance on herbal treatments, illustrating the persistent gap in formal health communication.

Psychosocial suffering was vividly illustrated in the field data, echoing findings by [13], who reported shame, anxiety, isolation, and internalized stigma among Ethiopian CL patients, especially women and children. These narratives align with the systematic review by [14], which frames stigma as multidimensional spanning enacted, internalized, and structural forms that severely impacts quality of life.

The gendered dimension of stigma emerges strongly. [15] systematically reviewed that women with CL face greater exclusion, particularly regarding marriageability and societal acceptance, underlining intersectional disparities. In our study, while both sexes experienced stigma, women bore the brunt filtered through cultural norms linking female value to appearance and purity.

Access to modern healthcare surfaced as a critical bottleneck, shaped by systemic inefficiencies and economic vulnerability. This mirrors findings by [16], identifying rising CL burden in Ethiopia post-civil unrest, with stretched health systems hampered by delays and limited resources. [17] further emphasize logistical obstacles drug shortages, inadequate infrastructure, and training gaps that limit decentralized CL care.

Despite these systemic challenges, Health Extension Workers (HEWs) and Community Advisory Groups (CAGs) play a hopeful but fragile role. [12] highlight HEWs as critical mediators of trusted biomedical knowledge, although they remain under-supported. [17] reinforce the promise of community-based interventions, tempered by questions of sustainability without proper systemic investment.

Finally, the economic and educational cost of CL is well-substantiated. [18] found that over 90% of Ethiopian CL patients experience moderate to severe impacts on schooling and work capacity due to lesion pain and stigma. These findings align with global frameworks identifying NTDs as entrenched poverty drivers, contributing to intergenerational disadvantage. In sum, recent studies from [12–18] strongly reinforce the multi-layered burdens of CL: entrenched cultural stigma, psychosocial distress, gender inequities, health system barriers, and economic hardship. Together, they call for integrated, culturally informed, gender-sensitive, and systems-strengthening responses to address CL not only as a disease, but as a phenomenon deeply embedded in social ecosystems.

## Conclusion

The ethnographic exploration of Cutaneous Leishmaniasis (CL) in affected Ethiopian communities reveals a layered and deeply human narrative one that extends far beyond the biomedical model. This study underscores how disease experiences are shaped not only by parasites and pathologies, but also by spiritual beliefs, gendered social roles, structural health system failures, and entrenched poverty. The persistence of supernatural and moral interpretations of CL combined with misconceptions about transmission demonstrates the enduring authority of culturally embedded explanatory models, which continue to operate in tension with biomedical messaging.

The psychosocial burden of CL emerged as a central theme, with stigma both enacted and internalized manifesting in social withdrawal, familial anxiety, and emotional isolation.

Particularly for women and children, CL imposed a disproportionate toll, echoing the gendered contours of health inequity found across other neglected tropical diseases. The implications of visible disfigurement reverberated across the social fabric, influencing marriageability, schooling, labor participation, and long-term life opportunities.

Systemic barriers to healthcare access characterized by logistical delays, economic strain, and treatment skepticism exacerbated the suffering. Despite the presence of frontline actors such as Health Extension Workers and Community Advisory Groups, these efforts remain under-resourced, operating within fragile institutional ecosystems that require sustained political and financial commitment. Yet their community-rooted strategies also demonstrate the value of participatory and culturally responsive approaches that go beyond top-down education models.

Ultimately, this study affirms that CL must be understood and addressed not only as a parasitic infection, but as a socially produced and sustained condition of marginalization.

Effective interventions must therefore be holistic, combining clinical treatment with robust health education, gender-sensitive policy design, and structural reforms that enhance system accessibility and equity. By attending to the voices of affected individuals and communities, particularly the most vulnerable, we may chart a path toward more inclusive and empathetic responses to CL and similar neglected diseases. The future of CL control lies not solely in therapeutic innovation, but in the ethical imperative to dismantle the social conditions that allow it to persist.

## Data availability

The qualitative data underlying this study including interview transcripts, field notes, and focus group summaries contain potentially identifiable and sensitive information and cannot be shared publicly due to ethical and confidentiality restrictions. Data are available upon reasonable request from the corresponding author and with approval from the Mekelle University Institutional Review Board (IRB) for researchers who meet the criteria for access to confidential data.

## Acknowledgments

The authors would like to thank the NIHR for supporting ECLIPSE. The authors would also like to acknowledge the three communities in three selected study sites for their support of research activities. Furthermore, we would like to recognise the ECLIPSE team for their assistance given throughout the project.

## Author contributions

Binega Haileselassie: Conceptualized and designed the study; conducted fieldwork including participant observation, interviews, and focus group discussions; led data analysis and interpretation; drafted the initial manuscript and coordinated subsequent revisions.

Zenawi Zerihun: Provided methodological guidance and critical input on ethnographic tools; contributed to data coding and analysis; reviewed and edited the manuscript for intellectual content and thematic coherence.

Afeworki Mulugeta: Supervised the overall research process; contributed to theoretical framing and contextual analysis; provided mentorship throughout the writing process and reviewed the final draft for academic rigor and policy relevance.

